## Supplementary material for "Solar UV–B/A Radiation is Highly Effective in Inactivating SARS-CoV-2": Supplemenatry Information

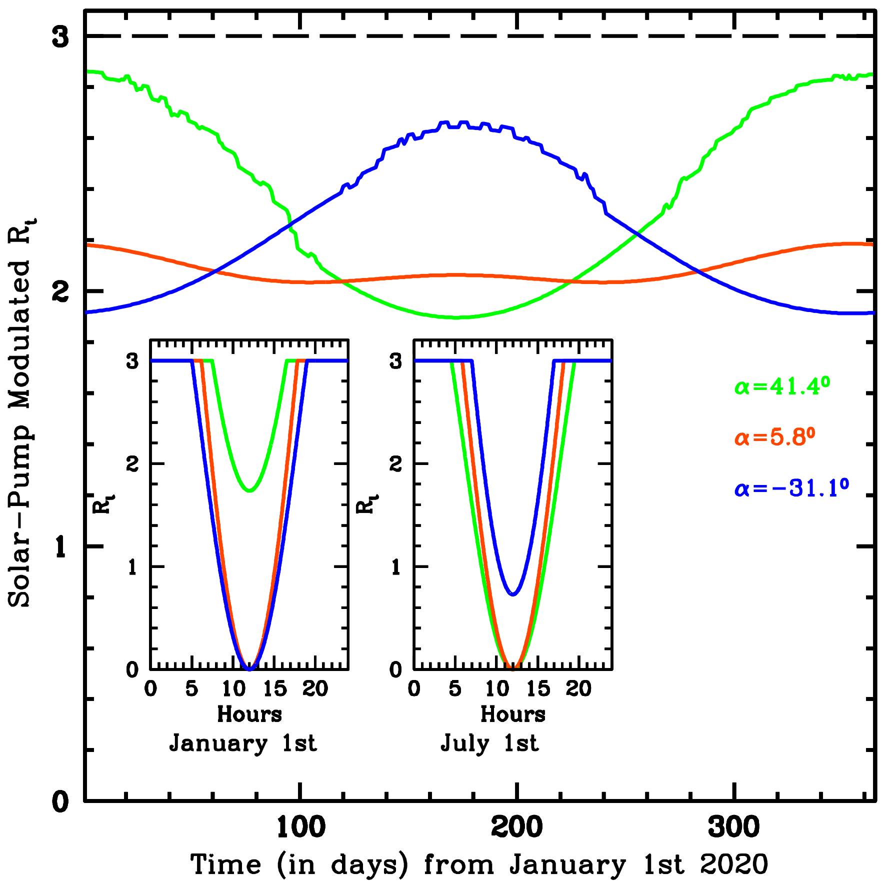


**Supplementary Figure S1 Main panel:** *day-by-day yearly modulation imprinted by the solar mechanism on the intrinsic reproduction number $R_0$=3 of the epidemic (black horizontal dashed line), at the three average latitudes of the countries of our N (green curves), T (orange curves) and S (blue curves) groups. Onsets: infra-day solar-pump modulation of $R_o$ at the three latitudes on January 1st (left onset) and July 1st (right onset) 2020, as labeled (see Methods for details).*

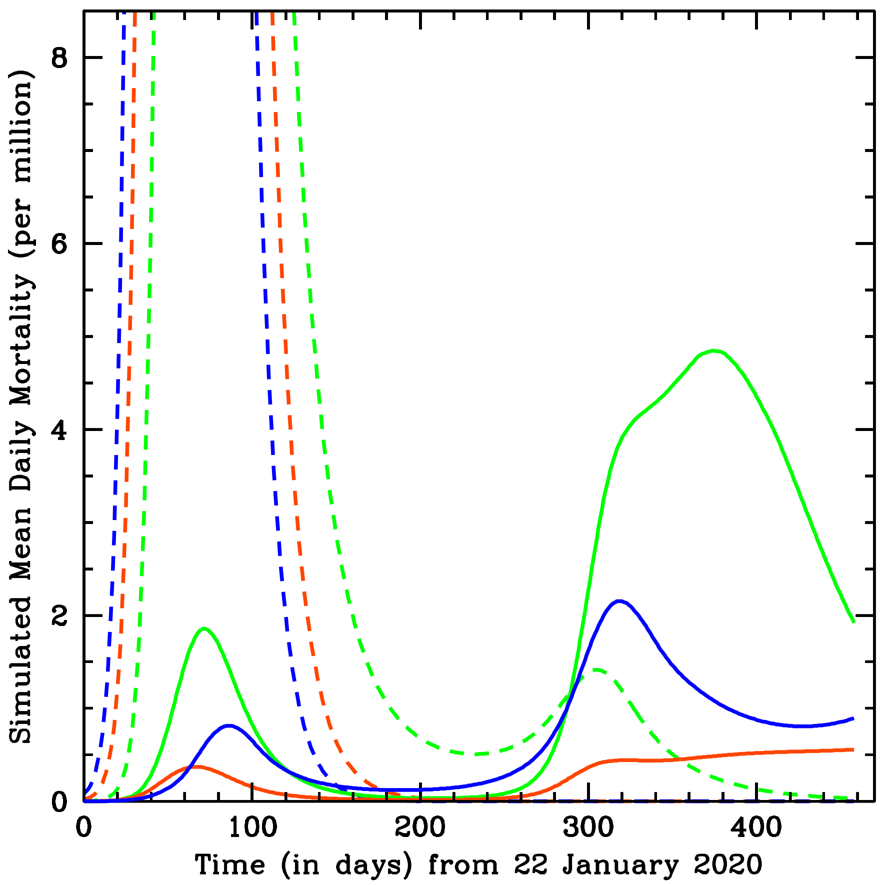


**Supplementary Figure S2 Simulated mean daily mortality curves as a function of time, from 22 January 2020 through April 2021,** ***with (solid curves) and without (dashed curves) the effect of the Solar-Pump (see Methods for details)*****.**
